# “A CTC Model Uncovers Metastatic Drivers and Prognostic Markers in Breast Cancer”

**DOI:** 10.1101/2025.01.16.25320597

**Authors:** Cristóbal Fernández-Santiago, Inés Martínez-Pena, Marta Paramés, Mario Rodríguez-Pérez, Hurtado Pablo, Carmen Abuín, Clotilde Costa, Ana Belén Dávila-Ibáñez, Rafael López- López, Roberto Piñeiro

**Author notes:** Correspondence: Roberto Piñeiro, Rafael López-López.

## Abstract

**Background:** Circulating tumor cells (CTCs) and CTC-clusters are pivotal in the metastatic process of breast cancer (BC). Owing to their low frequency, models replicating their biology should provide a robust platform for investigating the molecular mechanisms driving metastasis and identifying new biomarkers. We established and characterized a CTC-derived cell model from a mouse xenograft to explore its metastatic behavior and molecular profile, which allowed us to investigate the expression and prognostic significance of a set of genes associated with the metastatic potential of CTCs.

**Methods:** The CTC line (mCTC) derived from a MDA-MB-231 mouse xenografts was used in comparative functional analyses including cell cycle evaluation, colony formation, invasion, adhesion, and metastatic competency in zebrafish models. Transcriptomic profiling and functional assays were conducted to identify candidate genes and understand their roles in metastasis. Moreover, publicly available gene expression datasets of CTCs, CTC-clusters, and tumor tissue, from GEO and TCGA, were analyzed for the identification of a gene signature that was correlated with survival data. The signature was validated in an independent cohort.

**Results:** Compared with MDA-MB-231 cells, mCTC cells presented enhanced colony formation, invasion, and adhesion, and increased dissemination and survival in zebrafish. Transcriptomic analysis revealed that *SPARC* was significantly upregulated. Functional assays showed that *SPARC* overexpression was correlated with increased invasion and migration. Analysis of public datasets confirmed the high expression of *SPARC* in BC CTCs and CTC-clusters. Additionally, a 4-gene signature involving *SPARC*, *THBS1*, *VCL*, and *HSP90AB1* was identified that demonstrated strong prognostic value, predicting shorter overall and distant metastasis-free survival in the primary tumor setting. Validation cohorts confirmed its ability to distinguish high-risk patients. Elevated expression of the 4-gene signature in CTCs was also indicative of increased mortality risk.

**Conclusion:** mCTC exhibit distinct metastatic traits and molecular characteristics, highlighting a possible role of *SPARC* in CTC biology and its potential as a prognostic marker in BC metastasis. The identified 4-gene signature provides a robust prognostic tool for assessing patient risk and guiding therapeutic strategies. Further investigations into the mechanistic role of *SPARC* may reveal new therapeutic targets for managing BC progression.

## Introduction

Breast cancer (BC) is the leading cause of cancer-related death among women worldwide. Approximately 10% of newly diagnosed patients already have metastasis, and one-third will develop it within five years after diagnosis (1). Metastasis remains a major challenge in BC management, highlighting the urgent need for clinical strategies to treat metastatic BC (MBC) patients more effectively. A critical aspect of the metastatic process is the dissemination of circulating tumor cells (CTCs) from the primary tumor into the bloodstream, enabling the colonization of distant organs by these tumor cells. Notably, recent studies have highlighted the presence and potential significance of CTC-clusters, which are aggregates of CTCs that exhibit enhanced metastatic capacity compared with individual CTCs (2, 3).

CTCs offer significant prognostic and predictive value. The presence and quantity of CTCs and CTC-clusters in a patient’s blood can provide crucial insights into the aggressiveness of the disease and its potential for metastasis, thereby aiding in risk stratification and prognosis determination (2–8). Additionally, molecular and genetic analyses of CTCs can reveal specific mutations and alterations that are present in cancer, which can inform personalized treatment strategies and predict responses to targeted therapies. This makes CTCs a valuable tool in the management of cancer, enabling real-time monitoring of disease progression and treatment efficacy and facilitating adjustments to therapeutic approaches (9). But also, they are ideal sources for the identification of prognostic molecular biomarkers to help predict disease outcomes such as the likelihood of metastasis and overall survival rates.

CTCs are rare and highly heterogeneous and infrequent, complicating their isolation and characterization. Their presence in the bloodstream is a critical indicator of metastatic potential, so understanding their biology is key to advancing cancer treatment and prognosis. The development of cellular models that replicate the *in vivo* traits of CTCs and CTC-clusters is crucial for advancing our understanding of metastasis and improving therapeutic strategies (10, 11). Despite their clinical relevance, the biological characteristics and molecular mechanisms underlying the metastatic behavior of CTCs and CTC-clusters in MBC are not fully understood.

To address this gap, we established a novel CTC-derived cell model from a human triple-negative breast cancer (TNBC) mouse xenograft (12). This model aims to elucidate the functional properties and transcriptomic profile of these cells, providing a unique platform for investigating the metastatic traits of CTCs and identifying potential therapeutic targets.

In this study, we comprehensively characterized the CTC-derived culture (mCTC) in terms of its invasive and metastatic capabilities through *in vitro* functional assays, compared with parental MDA-MB-231 cells. Additionally, transcriptomic analysis was performed to identify differentially expressed genes (DEGs) and their potential roles in metastasis, revealing the significant upregulation of *SPARC*. SPARC, also known as osteonectin or BM-40, has been previously implicated in various cancers, demonstrating its ability to modulate the extracellular matrix (ECM) and influence tumor cell behavior (13–16). We therefore investigated the functional relevance of SPARC and evaluated its expression levels in CTCs and CTC-clusters from clinical samples through bioinformatics analysis of publicly available transcriptomic data. Lastly, we mined publicly available datasets to evaluate the expression of *SPARC* and related genes as prognostic biomarkers in early breast cancer.

## Materials and methods

### Cell culture, CTC isolation, and mCTC generation

The human triple-negative BC cell line MDA-MB-231, and the luminal A BC cell line MCF7, both expressing the enhanced green fluorescent protein (eGFP) and the luciferase (Luc) gene, were used for the experiments. MDA-MB-231 cells were purchased from Tebu-Bio (Barcelona, Spain), and MCF7 cells were purchased from GeneCopoeia, Inc (Rockville, MD, USA). In particular experiments, the CAF fibroblast line (17) was used as a control. The cells were cultured in high-glucose DMEM supplemented with 10% fetal bovine serum (FBS) and 1% v/v Penicillin/Streptomycin (P/S), at 37 °C in a humidified atmosphere containing 5% CO_2_, supplemented with Insulin + Transferrin + Selenium (GIBCO) for CAF culture. MDA-MB-231 cells. The mCTC cell line was established from CTCs and CTC-clusters isolated from the blood of mice injected with metastatic MDA-MB-231 cells as described previously (12). Briefly, CTCs isolated using the EasySep™ Mouse T-Cell Isolation Kit (StemCell Technologies), a negative selection method, were initially cultured in low-attachment 96-well plates (Corning) with 200 μL of Mammocult™ medium (StemCell Technologies), supplemented with hydrocortisone (0.48 μg/mL), heparin (4 μg/mL), bFGF (20 ng/mL), EGF (20 ng/mL), B27 supplement (4% v/v), progesterone (0.4 μg/mL), β-estradiol (0.4 μg/mL), ultraGRO™ (5% v/v), and penicillin/streptomycin (1% v/v). Cultures were maintained under hypoxic conditions (37 °C) for one week, with media renewal every two days. The cells were subsequently cultured under normoxic conditions (5% CO₂) at 37 °C, and a cell line referred to as mCTC was established. To generate MDA-MB-231-ULA, MDA-MB-231 cells were cultured in supplemented Mammocult™ medium under normoxic conditions.

Conditioned media were obtained by incubating the mCTC and MDA-MB-231 cells in DMEM for 3 days. After incubation, the medium was centrifuged and filtered with a 0.2 μm filter to remove cell debris.

### Cell cycle evaluation

Flow cytometry was conducted following propidium iodide (Sigma-Aldrich) staining of the cells to analyze the distribution of cells across different phases of the cell cycle, using a Cytek® Northern Lights-CLC cytometer (Cytek).

### Invasion assay

The invasive ability of mCTC cells was assessed via a Transwell® invasion assay (Corning, Madrid, Spain). The cells were seeded in serum-free medium and allowed to invade through growth factor reduced Matrigel-coated (Corning) Transwells in a chemoattractant-rich medium. After 24 hours, invading cells were fixed, stained with crystal violet, and quantified using a Leica DMi8 microscope (Leica Microsystems) and the free software ImageJ Fiji (NIH Image).

### Endothelial adhesion assay

Cell adhesion properties were evaluated by seeding them onto a monolayer of human umbilical vein endothelial cells (HUVECs) purchased from ATCC (USA). After a 45-minute incubation, non-adherent cells were washed off, and the number of cells adhering to the endothelial layer was quantified using a Leica DMi8 microscope and the free software ImageJ Fiji (NIH Image).

### Colony formation assay

Cells were suspended in 0.3% *w/v* molecular grade agarose (Fisher BioReagents), penicillin/streptomycin, and 10% FBS in DMEM and layered on top of a previously solidified 1.5 mL 0.5% *w/v* agarose, P/S, 10% FBS in DMEM in a 6-well multiwell plate. The cells were allowed to grow for a period of 3-4 weeks. Colonies growing in agarose were fixed and stained with crystal violet (Sigma-Aldrich, St. Louis, MO, USA) in methanol, for 1 hour at RT, with shaking. The number of colonies was counted and the area of each colony was determined using a Leica DMi8 microscopy.

### Hanging drop assay

MDA-MB-231 and mCTC cells (4×10^4^) were resuspended in either MammoCult or DMEM, and 20 μL drops (1×10^3^ cells/drop) were seeded onto the culture plate lids. Lids were rotated quickly and placed onto the cell culture plates. Cultures were kept in the incubator and pictures were taken at 48 and 72 hours to assess mammosphere formation.

### Wound healing assay

MCF7 cells were seeded in a 24 well plate as a monolayer and a wound was created. The culture medium was removed, the wells washed with PBS to eliminate dead cells, and the conditioned media was added. Images were obtained at 0, 24 and 48 hours using a Leica DMi8 microscope (Leica Microsystems), and the wound size was observed over time. Wound width was determined by measuring the area between the two parts of the monolayer in the same field of view at every time point with ImageJ Fiji (NIH Image).

### Metastatic competency in zebrafish models

Zebrafish husbandry, handling and experiments were performed as previously described (18). In brief, approximately 250 MDA-MB-231, MDA-MB-231-ULA or mCTC cells were injected into the perivitelline space of 48 hour postfertilization embryos using a microinjector IM300 (Narishige). Xenografted embryos were incubated for 3 days (120 hours post fertilization) and tumor dissemination and cell survival were assessed by fluorescence microscopy using a Leica DMi8 microscope acquiring images every 24 hours. For the experiments with small number of cells (mCTC single cells vs. mCTC small clusters), the cells were released into the pericardial space of the zebrafish embryo using a CellTram Oil micromanipulator (Eppendorf). Fluorescence images from zebrafish xenografts were analyzed using the free software Quantifish (Zebrafish Image Analyser). Quantification, statistical analysis and graphic representation were performed using the GraphPad Prism 8 software (GraphPad Software).

### Transcriptomic profiling by RNA-seq

RNA from the mCTC cell line and the MDA-MB-231-ULA populations was extracted using the RNeasy Micro kit (Qiagen) according to the manufacturer’s instructions and quantified in the Nanodrop OneC Microvolume UV-Vis Spectrophotometer (Fisher Scientific). RNA integrity was checked using the 4200 TapeStation System and TapeStation software (Agilent Technologies). The RNA was sequenced on the Illumina NovaSeq 6000 System, with 150 paired-end reads (2x 150 bp) and 100 M total reads. The expression profile was determined and expressed as FPKM (Fragment per Kilobase of transcript per Million mapped reads), RPKM (Reads Per Kilobase of transcript per Million mapped reads), and TPM (Transcripts Per Kilobase Million). Differentially expressed gene (DEG) analysis was performed using EdgeR, with the following cut-off: FC (fold change) ≥ 2 and p value < 0.05 for pairwise comparison.

### Gene and protein expression analysis

To validate the transcriptomic findings of the RNA-seq, RT-qPCR, Western blotting, and immunofluorescence were performed. For RT-qPCR, RNA was extracted from the cells using the High Pure RNA Isolation Kit (Roche). cDNA was synthesized using M-MLV Reverse Transcriptase (Invitrogen) and specific TaqMan probes used for SPARC (Hs00234160_m1), ITGB3 (Hs01001469_m1), COL6A3 (Hs00915125_m1), and TIMP3 (Hs00165949_m1). GAPDH (Hs99999905_m1) was used as an endogenous control gene. PCR reactions were conducted on a Roche LightCycler 480 II system (Roche). Relative gene expression levels were calculated using the ΔΔCt method. For Western blotting, protein was extracted using RIPA buffer, separated by SDS-PAGE, and transferred to PVDF membranes. The membranes were probed with antibodies against SPARC (AF941, R&D) and tubulin (#2146, Cell Signaling Technology), and detection was carried out using enhanced chemiluminescence (ECL; Thermo) using a ChemiDoc Imaging System (Bio-Rad). Band intensities were quantified using ImageJ software. For immunofluorescence, cells on coverslips were fixed with PFA 4% (w/v), permeabilized with InsidePerm (Miltenyi Biotec) and incubated with a primary antibody against SPARC (ON1-1, 33-5500, Invitrogen) and a secondary anti-mouse Alexa Fluor™ 647 antibody (Invitrogen). NucBlue™ Live ReadyProbes™ Reagent (Invitrogen) was used for DNA staining. Fluorescence images were acquired with a Leica DMi8 microscope (Leica Microsystems).

### Transcriptomic profiling and differential gene expression (DEG) identification

Transcriptomic data, encompassing both microarray and RNA-seq datasets, were obtained from the GEO database. Microarray data were processed and normalized using the Robust Multi-array Average (RMA) method via the rma() function from the affy package in R. This method includes background correction, quantile normalization, and probe-level summarization, which transform microarray intensities into log2 expression values. For the RNA-seq data, raw count matrices were obtained through GEO website, and normalization and variance stabilization were performed using the voom function of the limma package. The voom method transforms count data into log2 counts per million (logCPM) while accounting for the mean-variance relationship, enabling subsequent linear modeling with limma to assess differential expression. For datasets pooled from multiple experiments, batch effects were identified and corrected using the removeBatchEffect function in limma. For the GSE51827 and GSE86978 datasets, we adhered to the methodology initially employed by the original authors. This involved conducting an intrapatient comparison of CTC-clusters and single CTCs using the DEGseq package (version 1.56.1), with the analysis parameters set to the MARS method, a q-value threshold of 0.05, and a fold change criterion of 1.5.

### Public dataset analysis and gene signature identification

Publicly available datasets from GEO were analyzed to explore DEGs in CTCs vs healthy donors’ blood (GSE 148991). Additionally, single-cell RNA-seq data from paired individual CTCs and CTC-clusters were analyzed using GSE51827 and GSE86978 datasets. The prognostic performance of the 4-gene signature in CTCs (*SPARC*, *THBS1*, *VCL*, and *HSP90AB1*) was evaluated in two GEO datasets (GSE144494, GSE215886) containing OS data.

Data from the TCGA, which is publicly accessible, were analyzed to assess the overall survival (OS) and disease-free survival (DFS) associated with the gene signature derived from GEO dataset analysis of single CTCs vs CTC-clusters. Additionally, gene expression data were pooled from 11 GEO datasets (GSE11121, GSE1456, GSE16446, GSE17705, GSE19615, GSE2034, GSE20685, GSE4922, GSE5327, GSE6532, and GSE9195), with a focus on distant metastasis-free survival (DMFS).

### Network analysis

Network analysis was conducted to explore the interactions between DEGs, with protein-protein interaction (PPI) networks constructed using the STRING database (version 12.0). Key hub genes within the network were identified based on node degree and betweenness centrality, emphasizing genes linked to metastatic behavior. Survival analysis was performed using the survminer and survival packages in R. Patients were stratified into high-risk (CTCc-gene^High^) and low-risk (CTCc-gene^Low^) groups based on median gene expression levels.

### Immune and stromal microenvironment analysis

The xCell algorithm (https://xcell.ucsf.edu/) was used to estimate the relative abundance of immune cells and stromal cells for each patient sample by inputting mRNA expression matrices from the patients classified within the high and low risk groups according to the CTCc-gene signature. xCell can assess the level of infiltration of up to 64 cell types based on gene expression data.

### Statistical analysis

Data from functional *in vitro* assays were statistically analyzed using the Mann-Whitney test. Zebrafish experiments, when different time points were compared in one experimental group, were analyzed using the Wilcoxon test (non-parametric test for paired samples).

For gene expression and clinical data statistical analyses were performed using R (version 4.3.1). Descriptive statistics were calculated for baseline characteristics of study cohorts. Kaplan‒Meier survival curves were generated and compared using the log-rank test. Multivariate Cox proportional hazards models adjusted for clinical covariates were used to assess the prognostic significance of the 4-gene signature.

## Results

### 1.1. Functional characterization of a CTC-derived cell model (mCTC) from a TNBC mouse xenograft

The mCTC culture was previously derived from the CTCs and CTC-clusters isolated from the blood of mice orthotopically injected with metastatic MDA-MB-231 cells, using a negative enrichment technology (Supplementary Fig. S1, panel A) (see methods) (12). In these mice, CTCs and CTC-clusters were detected at a ratio of 1: 0.12 (CTC: CTC-cluster) (Supplementary Fig. S1, panel B). Cultured in ultra-low attachment (ULA) plates and specific cell culture media, mCTC grow in suspension mainly forming clusters and for an unlimited time (Supplementary Fig. S1, panel C). Cell cycle analysis of mCTC, MDA-MB-231 cells, and MDA-MB-231 cells cultured under the same conditions as mCTC (known as MDA-MB-231-ULA) was performed to evaluate their proliferation ability. The percentage of mCTC cells in the G0/G1, S, and G2/M phases was similar to that of parental MDA-MB-231, whereas MDA-MB-231-ULA presented signs of cell cycle arrest due to culture under non-adherent conditions (Supplementary Fig. S1, panel D).

Next, the performance of mCTC cells was compared with that of MDA-MB-231 cells via *in vitro* assays to evaluate colony formation, invasion, and adhesion to the endothelium. Compared with parental MDA-MB-231 cells, the mCTC line was able to efficiently form colonies in an anchorage-independent manner (*p* <0.05) and to invade in greater numbers in a transwell assay (*p* <0.01) (Fig. 1A, B). In addition, mCTC showed an enhanced capacity to adhere to a HUVEC monolayer (*p <*0.0001), but their ability to transmigrate through this monolayer was impaired (Fig. 1C, D). On the other hand, in a hanging drop assay, mCTC cells produced more and larger spheres than did MDA-MB-231 cells (Fig. 1E). Together, these results indicate that MDA-MB-231 cells that grew in a mouse and were isolated from the blood circulation acquired some functional traits that allowed them to behave differently from the parental cells initially injected into the mice.

**Figure 1.**
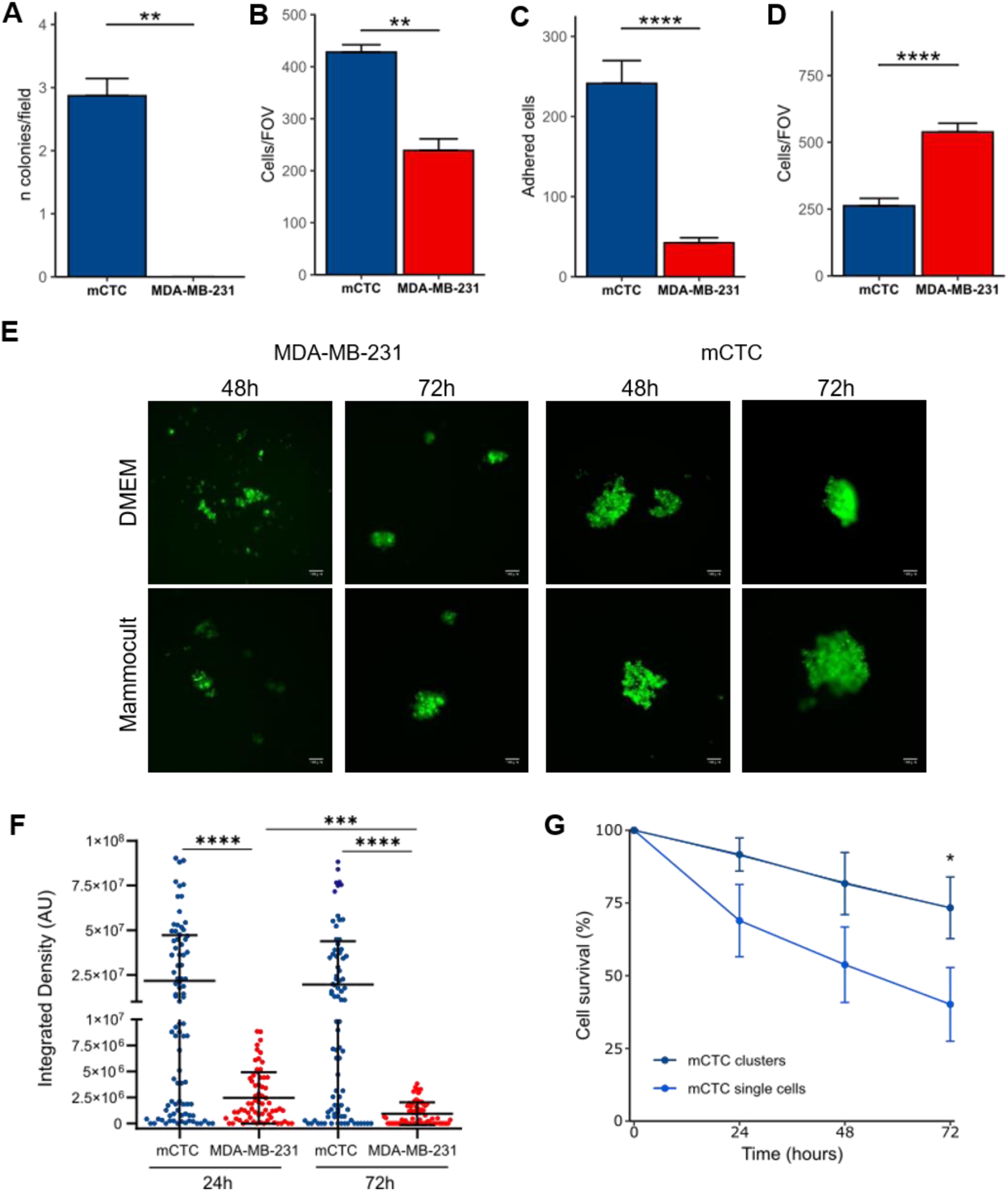
Evaluation of the metastatic competency of the mCTC cell line. **A** Number of colonies per field of view generated by mCTC and MDA-MB-231 cells in a soft agar colony formation assay (n= 3). **B** Invasion of mCTC and MDA-MB-231 cells (n= 3). The data are expressed as the number of invaded cells per field of view. **C** Number of mCTC and MDA-MB-231 cells adhered to a monolayer of HUVECs after 45 min of incubation. **D** Transmigration of mCTC and MDA-MB-231 cells represented as the number of migrated cells per field of view. **E** Representative images of the mammospheres formed by MDA-MB-231 and mCTC when incubated in DMEM or Mammocult media (scale bar 100 µm). **F** Fluorescence intensity of the mCTC cell line and the parental cell line MDA-MB-231 in the tail of the zebrafish at 24 and 72 hours post-injection (n= 6). **G** Survival of the mCTC clusters and individualized mCTC cells in the tails of the zebrafish. \**p <* 0.05, \*\**p <* 0.01, \*\*\**p <* 0.001, \*\*\*\**p <* 0.0001.

Moreover, we tested the dissemination and survival capacity of mCTC cells in a zebrafish assay of metastasis, as previously described (18). Equal numbers of mCTC or MDA-MB-231 cells were injected into the perivitelline space of the zebrafish embryo, and the presence of disseminated cells in the fish tails was determined 24 and 72 hours later on the basis of fluorescence intensity quantification. Compared with the parental MDA-MB-231 cells, the mCTC line presented enhanced metastatic behavior in the zebrafish model, with a greater number of disseminated cells in the tails 3 days post injection (mean integrated intensity of 1.96×10^7^ for mCTC vs. 9.32×10^5^ for MDA-MB-231; *p* <0.0001) and increased survival over time (Fig. 1F). These differences were also substantial compared with those of the MDA-MB-231-ULA cells (*p* <0.0001), as they disseminated and survived in much lower numbers (Supplementary Fig. S1, panel E). To further investigate the survival capacity of mCTC in circulation, small numbers of mCTC cells either as single cells or as small clusters were injected near the pericardial space of zebrafish embryos and followed on time (Supplementary Fig. S1, panel F). Compared with single mCTC cells, clusters of mCTC cells presented an increased resistance to cell death, with significant differences observed after 72 hours (*p* < 0.05) (Fig. 1G). While the number of single mCTC cells injected at 0 hours decreased at 24, 48, and 72 hours, this effect was much less pronounced in the clustered mCTC cells. This result indicates that the survival advantage of mCTC cells in circulation is partially lost when the clusters are broken up into individual cells. Taken together, these results suggest that mCTC line may represent a useful physiological CTC-cluster model, given its metastatic behavior, for investigating CTC-clusters biology.

### 1.2. Molecular profiling of mCTC cells and identification of SPARC

To gain better insight into the molecular changes acquired by mCTC cells a transcriptomic analysis was performed via RNA-seq. The mCTC gene expression profile was compared to that of MDA-MB-231-ULA cells (in culture for 4 days), to account for the potential modifications induced by the enriched cell culture medium and non-adherent conditions. RNA-seq data analysis revealed 1696 differentially expressed genes (DEGs) between mCTC and MDA-MB-231-ULA cells, with a fold change ≥2 and a p value < 0.05 (Fig. 2A). Specifically, 984 genes were significantly upregulated in the MDA-MB-231-ULA cells, whereas 712 genes were significantly upregulated in the mCTC cell line. To go deeper into the results, the DEG list was further analyzed with gProfiler for Gene Ontology (GO) analysis. This analysis revealed an enrichment of genes related to Biological Process (BP) such as the regulation of cell-cell adhesion, the regulation of cell adhesion, and extracellular matrix organization, and Cellular Components (CC) such as the collagen-containing extracellular matrix and cell-cell junction (Fig. 2B, and Supplementary Fig. S2, panel A).

**Figure 2.**
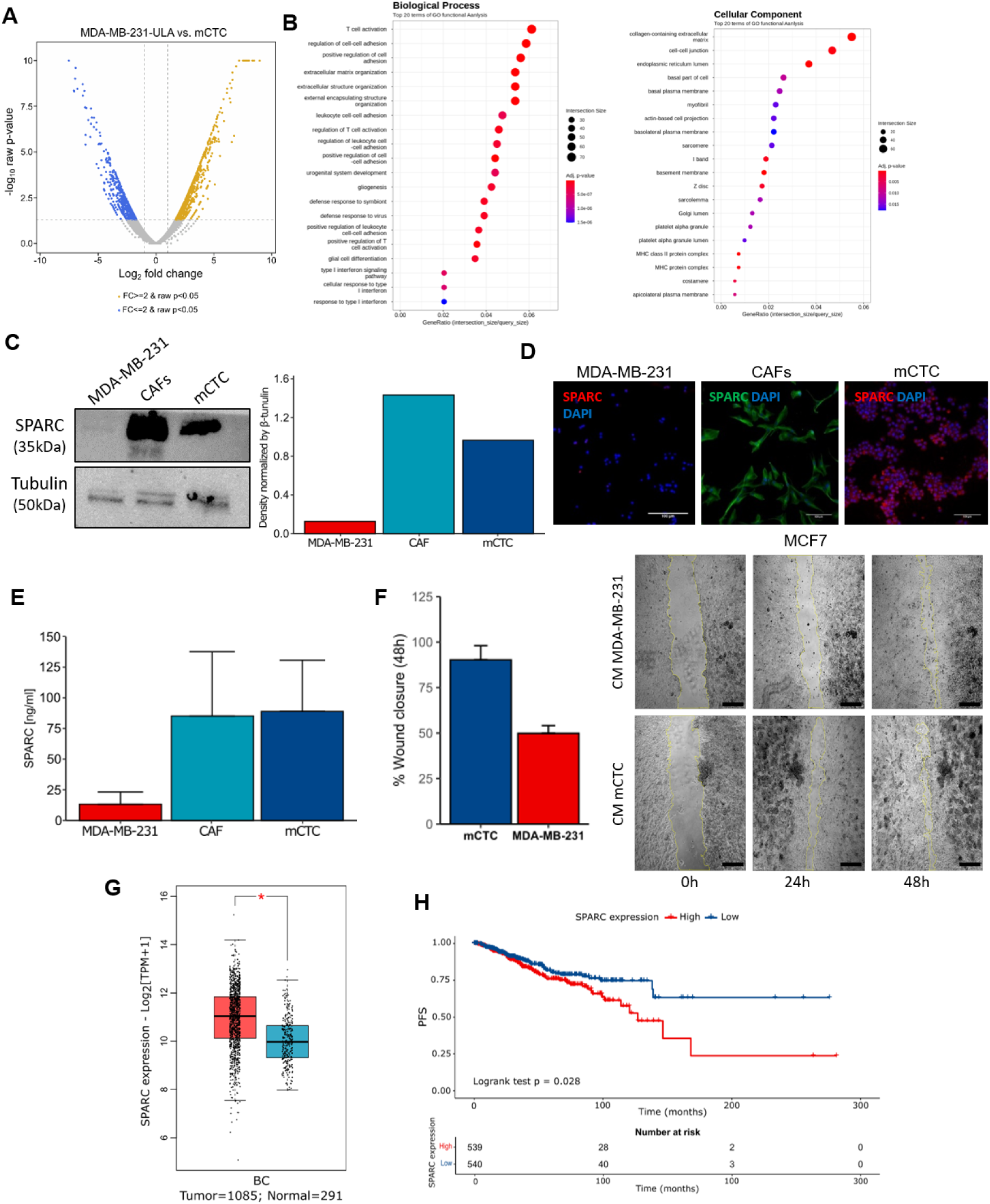
mCTC gene expression profiling by RNA-Seq and hit validation. **A** Volcano plot of the DEGs between mCTC and MDA-MB-231-ULA cells. **B** Gene Ontology (GO) enrichment analysis of DEGs showing the main terms related to (left) Biological Process and Cellular Components (right). **C** (left) Protein levels of SPARC in MDA-MB-231, CAF, and mCTC cells measured by Western blot and (right) densitometry (n=1). **D** Representative images of SPARC expression in MDA-MB-231, CAF and mCTC cells detected by immunofluorescence. Nuclei are labeled with DAPI (blue) (scale bar 100 µm). **E** Levels of SPARC detected in the culture media of the analyzed cells after 72 hours of culture (n=1, in duplicates). **F** (left) Quantification of the wound healing capacity of MCF7 cells (n= 2) in the presence of either mCTC or MDA-MB-231 conditioned medium (CM) and (right) representative images (scale bars 500 μm)**. G** mRNA expression level of SPARC in the BC TCGA database analyzed by GEPIA (\**p <* 0.05). **H** Kaplan–Meier plot of progression-free survival (PFS) analysis according to SPARC expression levels in the BRCA TCGA dataset. Breast cancer patients were divided into two subgroups according to high or low expression of SPARC using the median value as cut-off.

Among the top DEGs, the gene *SPARC*, encoding the “Secreted protein acidic and rich in cysteine” (aka. Osteonectin or BM-40) was highly upregulated in mCTC cells (Supplementary Table S1 shows the top 25 DEGs). This gene, together with 2 other upregulated genes (*ITGB3* and *COL6A3*) and 1 downregulated gene (*TIMP3*), — all genes related to remodeling and binding to the extracellular matrix — were selected for assessment of their gene expression levels by RT-qPCR for RNA-seq data validation. Data from mCTC, MDA-MB-231, and MDA-MB-231-ULA replicated the results of the RNA-seq analysis, showing an upregulation of *SPARC*, *ITGB3* and *COL6A3* in mCTC, and a downregulation of *TIMP3*. Total RNA from cancer-associated fibroblasts (CAFs), important producers of *SPARC* and *COL6A3*, was used as a positive control for expression (Supplementary Fig. S2, panel B).

*SPARC* has been previously shown to regulate cell migration and invasion, suggesting that it plays an important role in tumor progression and metastasis (19–21). We first demonstrated that SPARC protein expression was also upregulated in mCTC cells by western blot and immunofluorescence (Fig. 2C, D), and that mCTC cells were able to secrete SPARC into the cell culture medium to a similar extent to that of CAFs (Fig. 2E). These results suggest that some of the functional differences in invasion and migration displayed by mCTC could be related to the overexpression and secretion of SPARC. To test this hypothesis, we generated conditioned medium (CM) from both MDA-MB-231 and mCTC cells and incubated it with poorly invasive MCF7 cells in a wound healing assay. These experiments revealed that mCTC CM induced and enhanced the migration and wound closure of MCF7 cells compared with those of MDA-MB-231 CM (Fig. 2F). Thus, our data indicate that SPARC could play a key role in the molecular and functional characteristics of the mCTC cell line, and that its upregulation by mCTC cells may represent a specific adaptation of mCTC, potentially linked to their enhanced metastatic potential.

Importantly, GEPIA analysis (22) of gene expression data from 1085 breast tumor and 291 normal breast samples from the TCGA and GTEx projects showed that *SPARC* expression is increased in breast cancer tumor tissue (Fig. 2G). Similarly, SPARC is overexpressed in other tumor types, such as colon adenocarcinoma, head and neck squamous cell carcinoma, pancreatic adenocarcinoma, and melanoma (Supplementary Fig. S2, panel C). Moreover, survival analysis of the TCGA breast cancer dataset (n= 1079) revealed that patients with higher *SPARC* expression - using as a cut-off the median value of expression-in primary tumor tissue are at a greater risk of disease progression in terms of progression-free survival (PFS) (SPARC High *vs.* SPARC Low: HR_PFS_ 1.44 (95% CI 0.94–2.40), *p =* 0.028; Fig. 2B). However, no significant association was observed with overall survival (OS) (*p =* 0.34; Supplementary Fig. S2, panel D).

### 1.3. SPARC is highly expressed in BC CTCs and CTC-clusters

Very little information is available about the expression and role of *SPARC* in CTCs and CTC-clusters. A single CTC transcriptomic study in pancreatic ductal adenocarcinoma patients revealed that CTCs highly express *SPARC,* together with other stromal-derived extracellular matrix genes (23). These authors also showed that *SPARC* seems to be expressed by BC CTCs. Therefore, we aimed to determine the expression levels of *SPARC* in BC CTCs and CTC-clusters. We used publicly available datasets from the Gene Expression Omnibus (GEO) database. An analysis of RNA-seq data (GSE148991) comparing the gene expression profiles of CTC-derived short-term cultures from 6 patients and the cellular fraction of the buffy coat from 5 healthy donors showed that *SPARC* is highly upregulated in CTCs (logFC = 2.9, adj. *p* < 3.0×10^-4^; Fig. 3A). We next investigated the possible differences in expression between individual CTCs and CTC-clusters. We examined gene expression differences using two single-cell RNA-seq datasets (GSE51827 and GSE86978), including 65 individual CTCs and 36 CTC-clusters. DEG analysis identified 202 DEGs (Fig. 3B shows the top 25 upregulated genes in CTC-clusters). The GSE51827 dataset featured 10 patients with paired CTC-clusters and single CTCs, revealing that 6 of these patients exhibited upregulation of SPARC in CTC-clusters relative to single CTCs. The GSE86978 dataset, encompassing 22 patients, demonstrated a general upregulation of SPARC in CTC-clusters compared with single CTCs (Fig. 3C). This analysis reinforces the significant role of SPARC in CTC-cluster biology.

**Figure 3.**
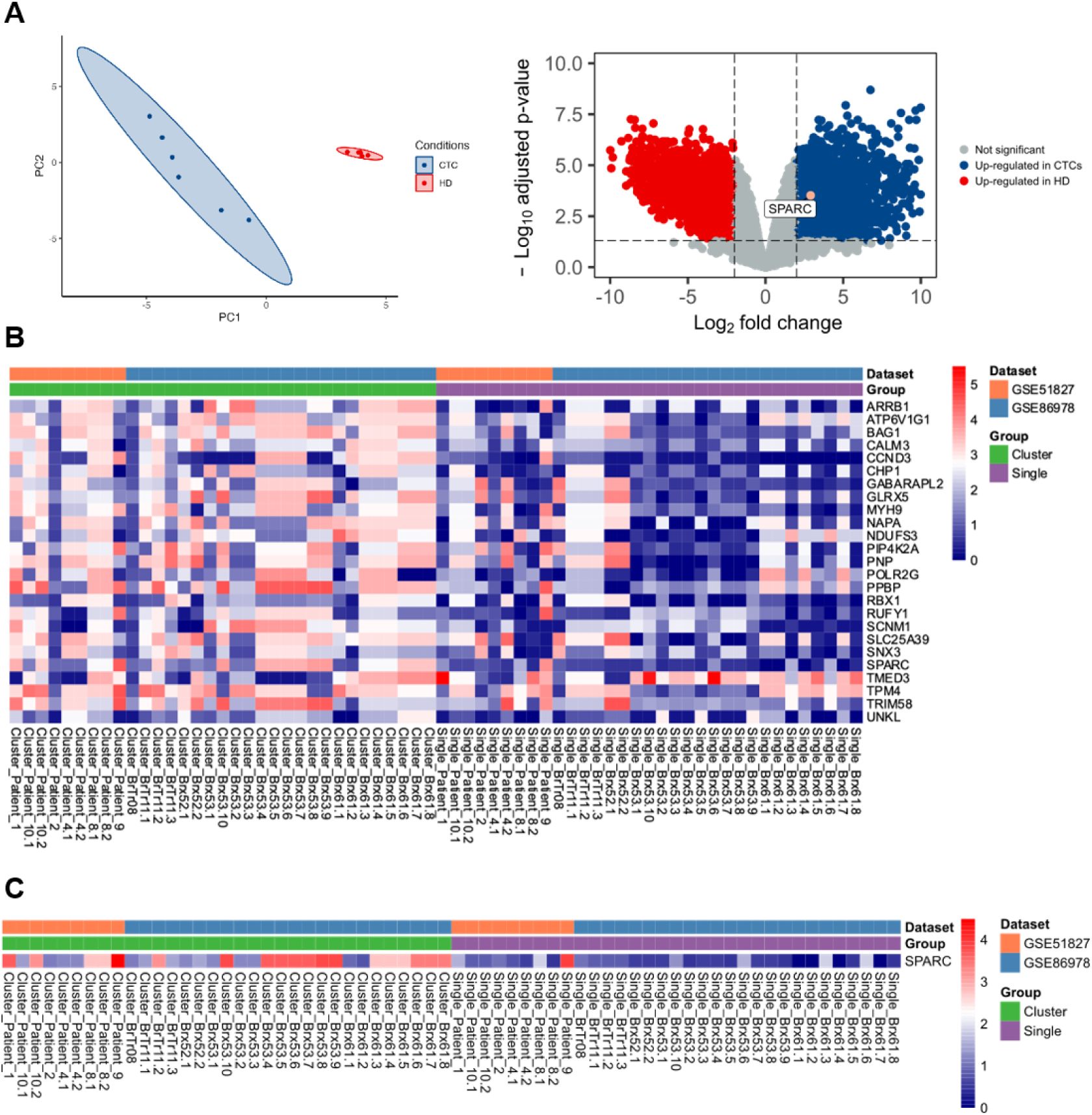
SPARC expression in BC CTCs and CTC-clusters. **A** (left) Principal component analysis (PCA) of mRNA gene expression in CTC cultures and the buffy coat of healthy donors (HD) and (right) a volcano plot showing the DEGs in the GSE148991 dataset. SPARC is labeled with a pink dot. **B** Heatmap showing the top 25 upregulated genes in CTC-clusters based on intrapatient comparison of CTC-clusters and single CTCs (GSE51827 and GSE86978, n= 65 single CTCs and n= 36 CTC-clusters), with a q-value < 0.05, and a log2 fold change ≥ 1.5. **C** Heatmap of SPARC mRNA expression in single CTCs *vs.* CTC-clusters, based on the intrapatient comparison shown in **B**.

### 1.4. Identification of a CTC-driven gene signature with prognostic value

To assess the potential functional interrelationships between *SPARC* and the commonly dysregulated genes in CTC-clusters, a protein‒protein interaction (PPI) network was constructed for the significantly enriched genes via the STRING database. A total of 14 nodes and 42 edges were identified within the PPI network, involving a core of 14 genes (*SPARC, VCL, ZYX, THBS1, TACSTD2, ANXA2, ANXA5, CDH1, CLU, CRIP1, CSRP1, HSP90AB1, ILK, ITGB5,* and *MGP*) (Fig. 4A, Supplementary Fig. S3, panel A). The functional enrichment analysis categorized the identified gene interactions into Biological Process (BP) and Molecular Function (MF) categories, providing speculative insights into their roles in CTC-cluster formation. Thus, in the BP category, focal adhesion, extracellular matrix organization, and adherens junctions were prominent, whereas in the MF category, cadherin binding was highlighted (Fig. 4B, C). The results suggest the potential involvement of these genes in CTC clustering and survival.

**Figure 4.**
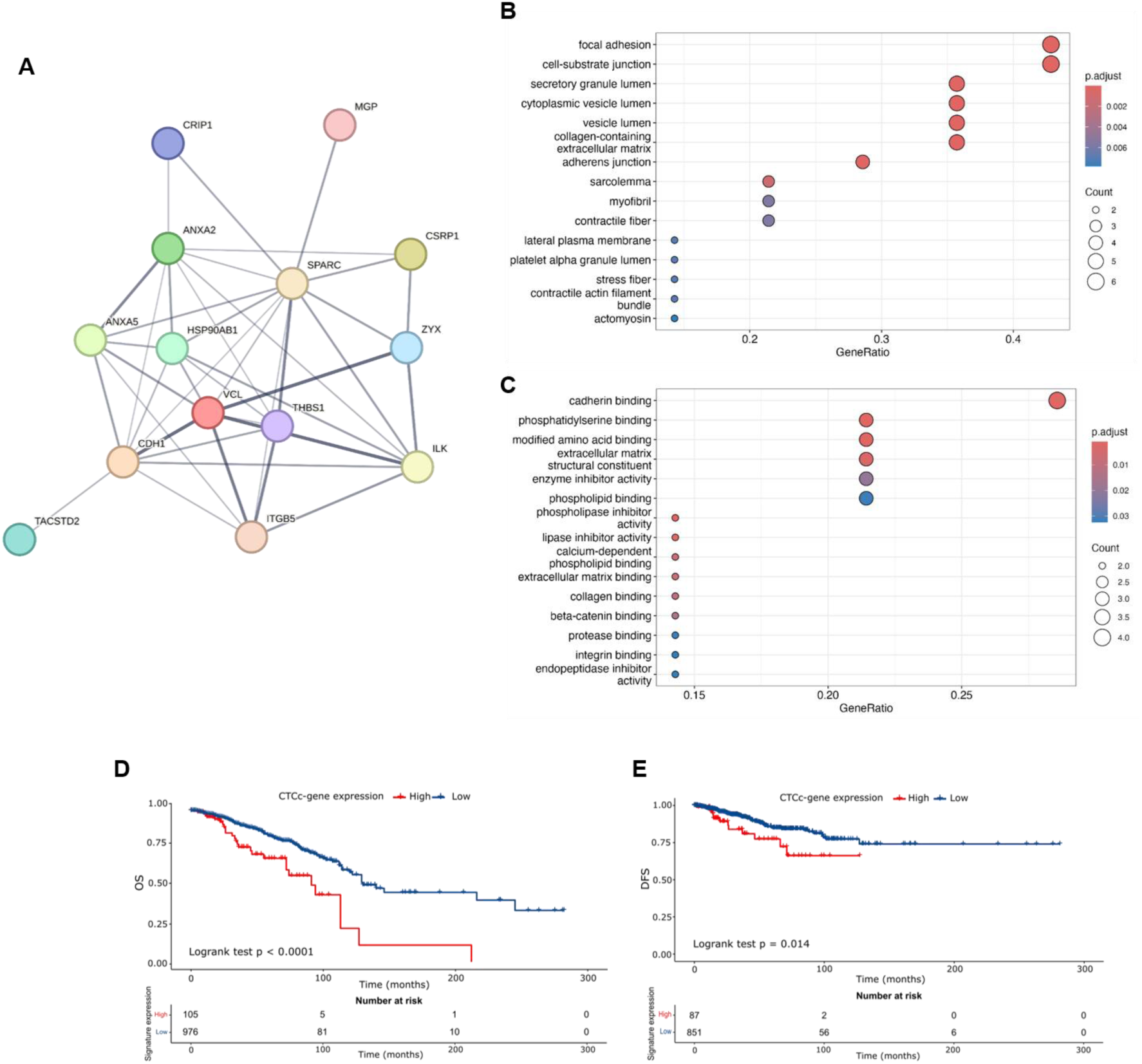
Development of a CTC-driven gene signature. **A** STRING analysis of genes commonly deregulated with SPARC in CTC-clusters. **B, C** GO enrichment analysis of genes commonly deregulated with SPARC in CTC-clusters showing the main terms related to BP and MF. Kaplan‒Meier plot for **D** overall survival (OS) and **E** disease-free survival (DFS) analysis according to expression levels of the 4-gene CTCc-gene signature in the primary tumors of the TCGA breast cancer dataset.

To delve into the prognostic implications of the identified CTC-cluster-derived genes (CTCc-gene) we used the TCGA breast cancer dataset, which contains primary tumor expression profiles. Our approach involved exploring combinations of two, three, and four CTCc-genes expressed in the tumor tissue that could predict patient outcomes. Patients were categorized into high-risk (CTCc-gene^High^) if each gene in the CTC cluster-derived signature showed higher expression than the median of all samples, and low-risk (CTCc-gene^Low^) if at least one gene was below this median. Ultimately, we identified a robust signature discriminating between risk groups comprising 4 genes: *SPARC*, *THBS1*, *VCL*, and *HSP90AB1*. Contrary to what was previously observed with *SPARC*, TCGA analysis revealed that none of these 3 genes are upregulated in breast cancer tissues compared with normal breast tissue (Supplementary Fig. S3, panel B) but that *THBS1* and *HSP90AB1* individually exhibited significant prognostic value for OS. Patients overexpressing *THBS1* and *HSP90AB1* had hazard ratios (HR) of 1.42 (95% CI: 1.03-1.95, *p =* 0.032) and 1.40 (95% CI: 1.02-1.93, *p* = 0.01), respectively (Supplementary Fig. S3, panel C). However, the synergistic effect of these four genes provided the most compelling prognostic indicator, identifying a group of high-risk patients (CTCc-gene^High^) with a heightened risk of death, with an HR of 2.348 (95% CI 1.545-3.567, *p <* 0.0001) for OS (Fig. 4D), and disease recurrence, with an HR of 1.36 (95% CI 0.94-2.40, *p =* 0.0159) for disease-free survival (DFS) (Fig. 4E).

### 1.5. The CTCc-gene signature predicts metastatic recurrence

To further validate our findings, we pooled the primary tumor gene expression data from 11 datasets within the GEO database (Table 1). The gene expression data were corrected for potential batch effects using limma package (Supplementary Fig. S4, panel A). This validation cohort consisted of 1645 patients, of which 593 patients had gene expression and OS data available. Stratifying patients on the basis of previously identified 4-gene signature revealed distinct prognostic groups in the OS analysis. Thus, the CTCc-gene^High^ group had an HR of 1.765 (95% CI 1.097-2.837, *p =* 0.018) with respect to the CTCc-gene^Low^ group (Fig. 5A).

**Figure 5.**
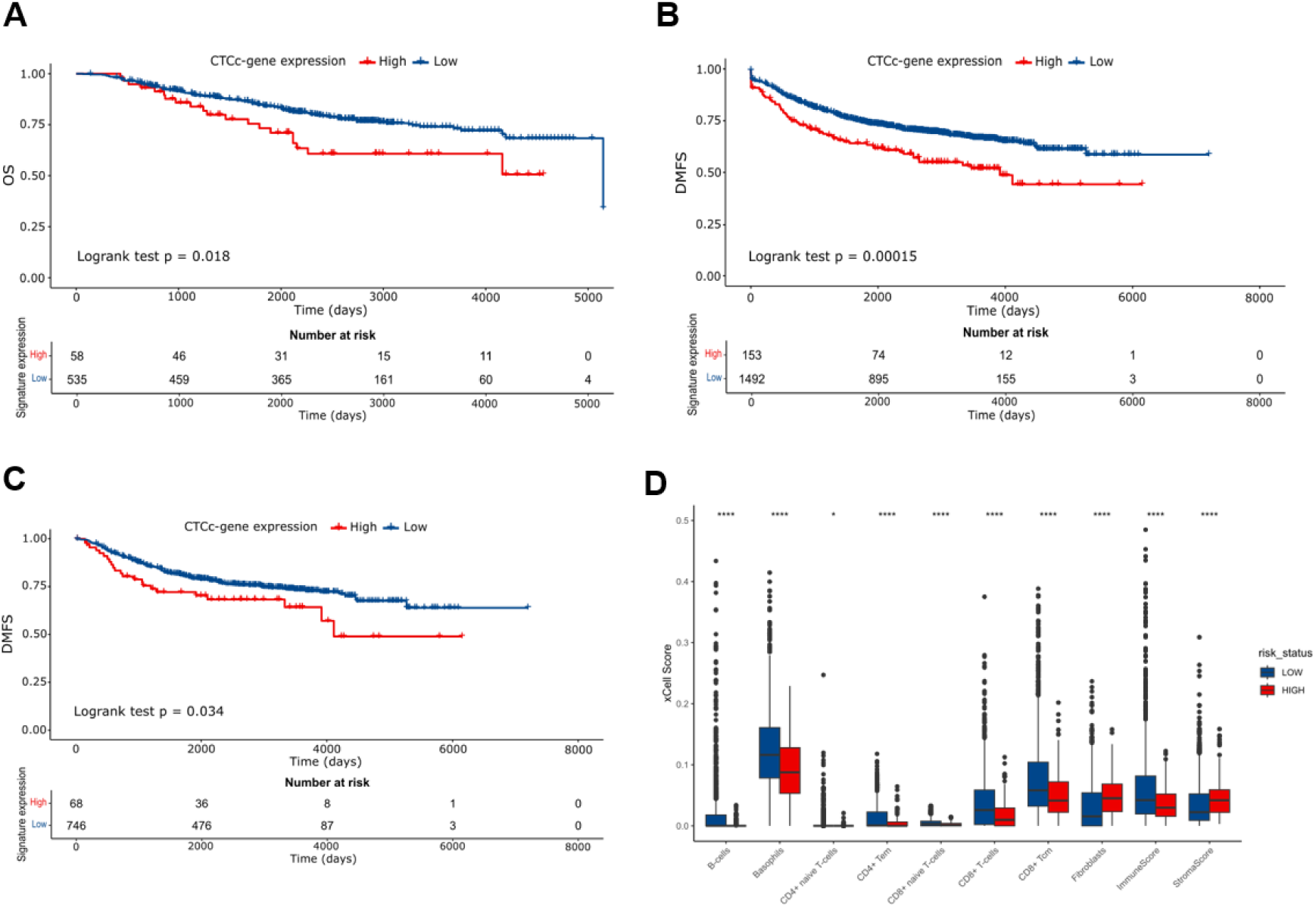
Validation of the prognostic value of the CTCc-gene signature in a pooled cohort of 11 datasets. **A** Kaplan‒Meier plot for OS analysis according to expression levels of the 4-gene CTCc-gene signature in the primary tumors of the validation cohort comprising 11 datasets from the GEO database (n= 593 patients). **B** Kaplan‒Meier plot for distant metastasis-free survival (DMFS) analysis according to expression levels of the 4-gene CTCc-gene signature in the primary tumors of the validation cohort (n= 1645 patients). **C** Kaplan‒Meier plot for DMFS analysis according to expression levels of the 4-gene CTCc-gene signature in the primary tumors of lymph node negative patients of the validation cohort (n= 814 patients). **D** Box plots showing the different infiltrating abundances of immune and stromal cells based on the classification of patient risk groups according to the CTCc-gene signature (xCell analysis).

We next wanted to test the value of the 4-gene signature in predicting the occurrence of distant metastasis. To this end, we took advantage of the available distant metastasis-free survival (DMFS) data (1645 patients) in the validation cohort. Survival analysis revealed that patients with a higher expression of the 4-gene signature in the primary tumor had a significantly shorter time to metastasis than patients with lower gene expression (CTCc-gene^High^ HR_DMFS_ 1.633 (95% CI 1.16-2.42), *p =* 0.00015; Fig. 5B). Interestingly, a subanalysis of the dataset within the group of patients without lymph node involvement, which is known to have a better prognosis than patients with lymph node positivity (24), showed the good prognostic performance of the 4-gene signature (HR_DMFS_ 1.592 (95% CI 1.032–2.457), *p =* 0.034; Fig. 5C).

To investigate the role of the tumor microenvironment in the differential metastatic potential of high-risk and low-risk patients, we conducted an xCell analysis (25) to assess the immune and stromal cell composition in primary tumor tissues from the pooled cohort of 11 GEO datasets. Patients were stratified on the basis of the expression of the identified 4-gene signature (CTCc-gene^High^ *vs.* CTCc-gene^Low^). Compared with their low-risk counterparts, high-risk patients presented significant decreases in the infiltration of CD8+ T cells, CD4+ T cells, and memory B cells, and increased infiltration of macrophages (Fig. 5D). The immune profile of these tumors suggests a less effective antitumor immune response. Concerning the stromal compartment, high-risk patients presented an increased presence of fibroblasts, indicating a potentially supportive niche for metastatic processes (Fig. 5D). Interestingly, these findings were supported by xCell analysis of the TCGA dataset, which also revealed similar differences in the immune composition of the tumors of patients from both risk groups (Supplementary Fig. S4, panel B).

### 1.6. The CTCc-gene signature in CTCs predicts OS

We have identified a set of genes enriched in CTC-clusters whose expression at the primary tumor site has prognostic value, identifying a high-risk group of patients with shorter OS and time to metastasis (DMFS). Therefore, we investigated the prognostic performance of the 4-gene signature in CTCs of two GEO datasets (GSE144494 and GSE215886) containing OS data. The combined higher expression of *SPARC*, *THBS1*, *VCL*, and *HSP90AB1* in CTCs exhibited significant prognostic value, allowing the identification of a group of patients with an increased risk of death. Indeed, Cox regression analysis of GSE144494 revealed that patients with CTCc-gene^High^ CTCs were at a greater risk of death than those with CTCc-gene^Low^ CTCs (HR_OS_ 10.020 (95% CI 1.070-93.808), *p =* 0.015; Fig. 6A). A similar, but not significant, result was obtained in the analysis of the GSE215886 dataset (CTCc-gene^High^ group HR_OS_ of 6.552 (95% CI 0.719-59.740), *p =* 0.057; Fig. 6B).

**Figure 6.**
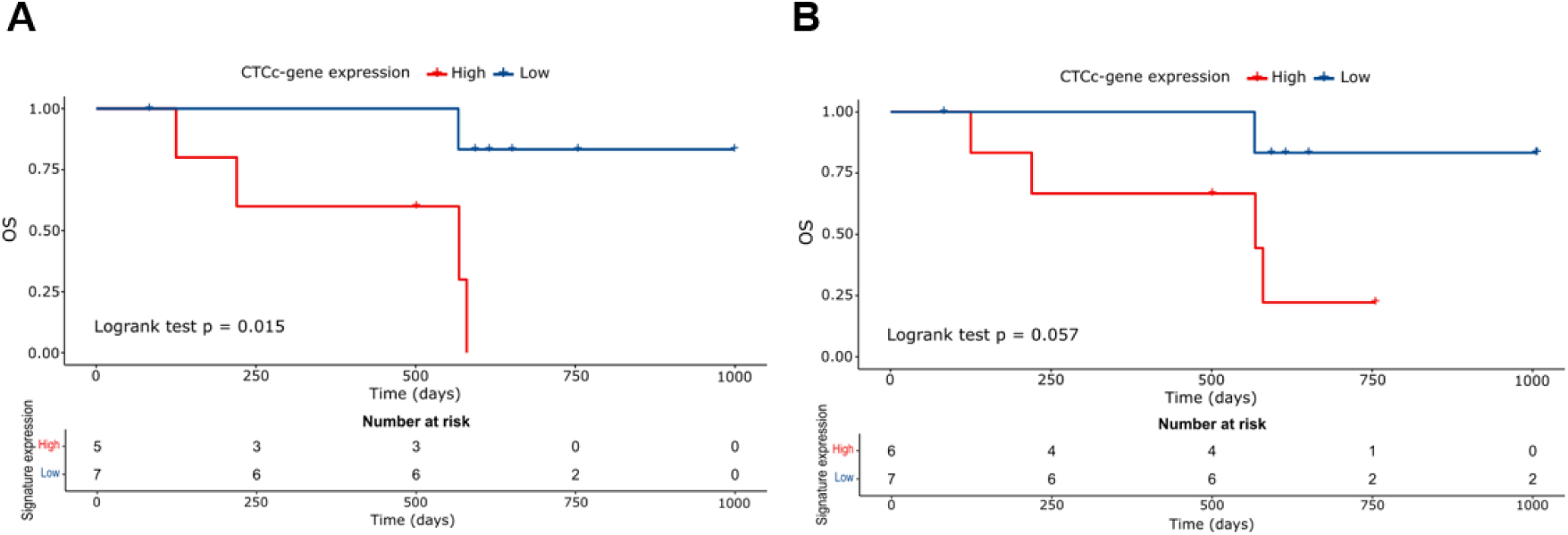
Prognostic performance of the CTCc-gene signature in CTC dataset. Kaplan-Meier plot of OS analysis according to expression levels of the 4-gene CTCc-gene signature in CTCs from the **A** GSE144494 and **B** GSE215886 datasets.

## Discussion

The results of this study suggest a role for *SPARC* in the metastatic behavior of CTCs and CTC-clusters in breast cancer. By establishing and characterizing a CTC-derived cell model (mCTC) from a TNBC mouse xenograft, resembling of CTC-clusters, we have gained valuable insights into the molecular and functional properties that contribute to the aggressive nature of these cells, suggesting that this model represents a valuable tool for future research into the mechanisms of CTC-cluster mediated metastasis.

The differences observed between mCTC cells and parental MDA-MB-231 cells highlight the unique characteristics of these cells. Compared with parental MDA-MB-231 cells, mCTC cells exhibited significantly greater colony formation, invasion, and adhesion to the endothelium. These observations were corroborated by *in vivo* experiments using zebrafish models, where mCTC cells demonstrated increased metastatic competency and survival. Moreover, the functional assays conducted in this study suggest that *SPARC* seems to be associated with increased colony formation and cell migration, and invasion capabilities. Moreover, the ability of mCTC-conditioned medium to induce higher wound closure and invasion in MCF7 cells than that of MDA-MB-231-conditioned medium further proves the potential role of SPARC in promoting invasive phenotypes. Thus, the increased expression of SPARC in these cells likely contributes to their enhanced metastatic behavior by facilitating the cellular processes critical for invasion and migration.

Our findings support the hypothesis that *SPARC* plays a critical role in BC metastasis. Previous studies have implicated SPARC in various cancers, demonstrating its ability to modulate the ECM and influence tumor cell behavior (13, 16, 26). In our model, SPARC appears to facilitate the detachment of tumor cells from the primary site, promote their survival in the bloodstream, and enhance their colonization of distant organs. These results align with the literature suggesting that SPARC interacts with integrins and other ECM components, promoting epithelial‒mesenchymal transition (EMT) and enhancing metastasis (19, 27, 28). Therefore, the increased metastatic potential of mCTC cells, reminiscent of that of the one of CTC-clusters, underscores the importance of targeting CTCs and CTC-clusters in therapeutic strategies. The broader implications of these findings are also supported by GEPIA analysis, which revealed that *SPARC* overexpression is limited not only to BC but also to other cancer types. In addition, survival analysis of TCGA breast cancer data suggested that higher *SPARC* expression is correlated with poorer progression-free survival (PFS) outcome, demonstrating that *SPARC* could be a potential prognostic biomarker in BC.

Our analysis revealed that the expression of *SPARC* in BC CTCs is upregulated compared with that in leukocytes, present in the cellular fraction of the buffy coat. Similarly, a previous study in pancreatic ductal adenocarcinoma revealed increased expression of *SPARC* in CTCs, along with other stromal‒derived extracellular matrix genes, whose knockdown in cancer cells suppresses cell migration and invasion (23). Furthermore, transcriptomic data analysis of CTCs and CTC-clusters isolated from clinical samples revealed altered expression of *SPARC*, which was consistently greater in CTC-clusters across the two datasets analyzed. Together, the results suggest that SPARC may play a crucial role in the biology of CTC-clusters, and that the elevated expression of *SPARC* in CTC-clusters indicates that SPARC could be a key mediator in facilitating the collective migration and invasion capabilities of these clusters, contributing to their increased metastatic competency. The identification of *SPARC* as a differentially expressed gene in CTC-clusters provides the rational to be considered a potential target for therapeutic intervention aimed at disrupting the metastatic cascade. In this regard, SPARC has been already postulated as a therapeutic target in the primary tumor (29). Moreover, these findings warrant further investigation into the mechanistic role of SPARC in CTC-clusters, which could increase our understanding into the molecular underpinnings of metastasis and inform the development of strategies to prevent the spread of BC. A caveat to these speculations is represented by previous reports suggesting that platelets coating CTCs could be a source of expression of mesenchymal markers, such as SPARC (2, 30). Nevertheless, mounting evidence suggests that the aggregation of platelets to CTCs promotes metastasis (31, 32).

On the other hand, our bioinformatic analysis of transcriptomic data from individual CTCs and CTC-clusters has provided valuable insights into the molecular differences that may drive metastasis in BC. The selected datasets, comprising a total of 65 individual CTCs and 36 CTC clusters, allowed for a comprehensive comparison of gene expression profiles between these two CTC populations, revealing a set of genes associated with *SPARC* and commonly upregulated in the CTC-cluster. Functional Enrichment Analysis revealed their involvement in biological processes such as focal adhesion, extracellular matrix organization, and adherens junctions. These processes may play crucial roles in enhancing the clustering ability and metastatic potential of CTCs (33–35). Genes such as *VCL* and *ILK*, which play roles in the formation and regulation of focal adhesions, may facilitate strong interactions between CTCs and surrounding stromal cells, providing the structural support necessary for cluster formation and migration. Extracellular matrix organization, influenced by proteins such as SPARC and THBS1, could create a favorable environment that supports CTC survival and proliferation. Furthermore, genes associated with adherens junctions and the extracellular matrix are speculated to be vital for maintaining CTC-cluster integrity, as they play significant roles in cell-cell junctions, which are potentially essential for the cohesion and stability of CTC-clusters during their circulation in the bloodstream. Moreover, cadherin binding has been revealed as an important molecular function, as it is crucial for cell-cell adhesion. It is speculated that cell-cell interactions are essential for maintaining the structural integrity of CTC-clusters and facilitating intercellular communication within CTCs (2, 36). By forming strong intercellular connections, cadherin binding may help CTCs resist shear stress in the bloodstream and promote CTC survival and metastatic dissemination. The identified hub genes within the PPI network suggest a complex interplay of cellular mechanisms that might support the formation and maintenance of CTC-clusters, all of which could contribute to the metastatic potential of CTC-clusters. Hence, understanding these interactions may allow us to identify potential therapeutic targets for disrupting CTC-cluster formation.

We identified a 4-gene signature (*SPARC*, *THBS1*, *VCL*, and *HSP90AB1*) that provides a novel prognostic marker in BC. Our study leveraged the comprehensive TCGA dataset to explore combinations of genes predictive of patient outcomes. By stratifying patients into high-risk (CTCc-gene^High^) and low-risk (CTCc-gene^Low^) groups, we identified a signature that consistently demonstrated prognostic value. In contrast to *SPARC*, TCGA analysis indicated that none of the three genes included in the signature (*THBS1*, *VCL*, and *HSP90AB1*) are upregulated in BC tissues compared with normal breast tissues. This discrepancy suggests a role for these genes in breast cancer that may not be solely defined by their expression levels in the tumor but rather by their collective impact on prognosis when considered in combination with *SPARC*.

At the prognostic level, *THBS1* and *HSP90AB1* individually emerged as notable predictors of OS within the TCGA cohort. However, the synergistic effect of *SPARC*, *THBS1*, *VCL*, and *HSP90AB1* had the strongest prognostic power. The CTCc-gene^High^ patient group, characterized by the simultaneous high expression of this 4-gene signature at the primary tumor, revealed a significantly increased risk of death (HR_OS_ of 2.348) and disease progression (HR_DFS_ of 1.36). Importantly, these associations persisted after adjusting for various clinicopathological variables in multivariate analysis, reinforcing the robustness of this gene signature in predicting patient outcomes. Further validation of the signature in a new cohort comprising 593 patients from multiple GEO datasets confirmed its ability to discriminate patients into distinct prognostic groups based on OS analysis, which maintained statistical significance in multivariate Cox regression analysis, further validating its utility as an independent prognostic tool.

Breast cancer is a chronic disease, meaning that late recurrence and metastasis are significant concerns for long-term patient outcomes. These late events can arise many years after the initial diagnosis and treatment, representing a persistent risk that needs ongoing vigilance and management throughout patients’ lives. The potential clinical utility of this signature comes from its ability to predict shorter DMFS, identifying a group of patients at greater risk of metastatic recurrence. Our analysis revealed that the 4-gene signature maintains its prognostic value in this subgroup of patients, indicating that patients categorized as CTCc-gene^High^ had a significantly greater risk of developing distant metastasis than those in the CTCc-gene^Low^ group. This suggests that the 4-gene signature could be capturing molecular features associated with enhanced metastatic propensity, such as increased cell adhesion, migration, and survival abilities of tumor cells. Consequently, the 4-gene signature could serve to identify within this lower-risk cohort, those individuals who may benefit from more intensive monitoring and potentially adjuvant therapies.

Our analysis of the cellular composition of the primary tumors in the risk group patients suggests a lower degree of immune infiltration in the CTCc-gene^High^ group. The reduction in CD8+ T cells and memory B cells, key immune cell populations, suggests a less effective anti-tumor immune response, which may allow the tumors to evade immune surveillance and contribute to the increased metastatic risk observed in these patients. CD8+ T cells play a crucial role in directly targeting and eliminating tumor cells, and their diminished presence could impair immune control over tumor progression (37). Similarly, lower levels of memory B cells might result in less robust long-term immune surveillance (38), further compromising the immune system’s ability to prevent metastasis. On the other hand, tumors from high-risk patients exhibited a significantly increased infiltration of macrophages, suggesting a potentially more pro-tumorigenic microenvironment. Macrophages, particularly tumor-associated macrophages (TAMs), have been implicated in various pro-metastatic processes, including the enhancement of EMT, support of angiogenesis, and establishment of pre-metastatic niches (39–41). Therefore, although the exact mechanisms remain to be fully elucidated, the immune and stromal cell composition determined by the 4-gene signature aligns with the more aggressive clinical outcomes associated with high-risk patients, as evidenced by their shorter DMFS, and would allow us to speculate that these factors may facilitate the survival and dissemination of CTCs and CTC-clusters, potentially leading to the observed increase in metastatic spread in high-risk patients.

Lastly, the observed enrichment of *SPARC*, *THBS1*, *VCL*, and *HSP90AB1* in BC CTCs and their association with poorer survival outcomes could underscores the prognostic importance of the 4-gene signature not only in primary tumors but also in CTCs. The heightened expression in CTCs of these genes involved in critical processes such as cell adhesion, migration, and interactions with the extracellular matrix, might suggest that these cells possess enhanced capabilities to circulate within the bloodstream, evade immune surveillance, and ultimately establish metastatic lesions. From a clinical standpoint, it implies that patients whose CTCs express elevated levels of these genes may have a higher likelihood of disease progression and poorer treatment outcomes. This speculation raises interesting questions about the potential utility of these genes in monitoring disease progression and as predictive biomarkers for identifying patients at higher risk of metastatic relapse or therapeutic resistance as previously described.

Taken together, these findings highlight the potential of *SPARC*, *THBS1*, *VCL*, and *HSP90AB1* as potential biomarkers in BC prognosis. Their collective prognostic impact, validated across multiple datasets, underscores the robustness of this gene signature and highlights its clinical relevance and potential for guiding personalized treatment strategies.

While the results are promising, it is essential to interpret these speculations with caution, as there are limitations to consider. For instance, with regard to DMFS prediction in lymph node-negative patients as well as OS prediction based on the identification of high-risk patients by CTC gene expression, prospective validation in larger and independent cohorts is needed to confirm these findings. Additionally, further exploration of the biological function and interactions of these genes could provide deeper insights into their roles in breast cancer progression and metastasis.

## Ethics approval and consent to participate

Not applicable.

## Consent for publication

Not applicable.

## Author contributions

C.F.-S. was involved in the methodology, data analysis, interpretation and discussion, and original draft preparation. I.M.-P., M.P., M.R.-P., and P.H. were involved in the methodology and data analysis and interpretation. C.A. was involved in the methodology. C.C. and A.B.D.-I. were involved in the interpretation and discussion of the data. R.L.-L. was involved in the design and conceptualization of the study, interpretation of the data and drafting and revision of the manuscript. R.P. was involved in the design and conceptualization of the study, supervision, data analysis, interpretation of the data, and drafting and revision of the manuscript.

## Availability of data and materials

The datasets generated and/or analysed during the current study available from the corresponding author on reasonable request.

## Data Availability

All data produced in the present study are available upon reasonable request to the authors

## Acknowledgments

We thank Jose Antonio Trillo Franco for his technical assistance with the *in vitro* experiments.

## Abbreviations

BC: Breast cancer
BP: Biological Process
CAFs: Cancer-associated fibroblasts
CM: conditioned medium
CRC: Colorectal cancer
CTCs: Circulating tumor cells
DEGs: Differentially expressed genes
DFS: Disease-free survival
DMFS: Distant metastasis-free survival
ECM: Extracellular matrix
eGFP: Enhanced green fluorescent protein
EMT: Epithelial-to-mesenchymal transition
GEO: Gene Expression Omnibus
GTEx: Genotype-Tissue Expression
GO: Gene ontology
GEPIA: Gene expression profiling interactive analysis platform
HR: Hazard ratio
HSP90AB1: Heat Shock Protein 90 Alpha Family Class B Member 1
HUVEC: Human Umbilical Vein Endothelial Cells
MBC: Metastatic Breast Cancer
MP: Molecular Function
OS: Overall survival
PFS: Progression-free survival
PPI: Protein-protein interaction
SPARC: Secreted Protein Acidic and Cysteine Rich
STRING: Search Tool for the Retrieval of Interacting Genes/Proteins
TCGA: The Cancer Genome Atlas
THBS1: Thrombospondin-1
TNBC: Triple negative breast cancer
VCL: Vinculin

