## Supplementary material for "“A CTC Model Uncovers Metastatic Drivers and Prognostic Markers in Breast Cancer”": Supplemetary Table & Figures

### Supplementary information

### Supplementary Table S1. Top 25 DEGs between MDA-MB-231-ULA and mCTC cells.

| Gene_ID | Gene Symbol | Description | FC (231ULA/mCTC) | P-val |
|---|---|---|---|---|
| 3434 | IFIT1 | interferon induced protein with tetratricopeptide repeats 1 | 257,72 | 7,83E-09 |
| 3868 | KRT16 | keratin 16 | 491,6 | 7,83E-09 |
| 126353 | MISP | mitotic spindle positioning | 491,14 | 7,83E-09 |
| 6947 | TCN1 | transcobalamin 1 | 318,47 | 3,92E-08 |
| 9023 | CH25H | cholesterol 25-hydroxylase | 209,88 | 3,92E-08 |
| 6653 | SORL1 | sortilin related receptor 1 | 219,36 | 4,31E-08 |
| 6678 | SPARC | secreted protein acidic and cysteine rich | -183,92 | 4,96E-08 |
| 3872 | KRT17 | keratin 17 | 193,79 | 7,31E-08 |
| 1953 | MEGF6 | multiple EGF like domains 6 | 182,24 | 8,32E-08 |
| 8638 | OASL | 2'-5'-oligoadenylate synthetase like | 143,52 | 8,79E-08 |
| 91543 | RSAD2 | radical S-adenosyl methionine domain containing 2 | 233,58 | 1,17E-07 |
| 283120 | H19 | H19 imprinted maternally expressed transcript | 351,89 | 1,63E-07 |
| 3627 | CXCL10 | C-X-C motif chemokine ligand 10 | 121,21 | 3,13E-07 |
| 11082 | ESM1 | endothelial cell specific molecule 1 | -123,49 | 3,49E-07 |
| 9636 | ISG15 | ISG15 ubiquitin like modifier | 97,11 | 5,88E-07 |
| 284266 | SIGLEC15 | sialic acid binding Ig like lectin 15 | -126,36 | 1,20E-06 |
| 1907 | EDN2 | endothelin 2 | 97,62 | 1,31E-06 |
| 1917 | EEF1A2 | eukaryotic translation elongation factor 1 alpha 2 | 83,35 | 1,92E-06 |
| 4855 | NOTCH4 | notch receptor 4 | -80,46 | 2,05E-06 |
| 154064 | RAET1L | retinoic acid early transcript 1L | 91,47 | 3,09E-06 |
| 2358 | FPR2 | formyl peptide receptor 2 | -110,38 | 3,14E-06 |
| 3433 | IFIT2 | interferon induced protein with tetratricopeptide repeats 2 | 69,89 | 3,14E-06 |
| 4101 | MAGEA2 | MAGE family member A2 | -137,04 | 3,72E-06 |
| 3772 | KCNJ15 | potassium inwardly rectifying channel subfamily J member 15 | -72,94 | 3,81E-06 |
| 266740 | MAGEA2B | MAGE family member A2B | -134,61 | 3,83E-06 |

**Supplementary Figuress.**

**Supplementary Figure 1. mCTC establishment and functional assays. A** Schematic representation of the workflow followed for the establishment of the mCTC cell line. **B** CTCs and CTC-clusters are detected in the blood of mice in a proportion 1:0,12 (CTC:CTC-cluster). **C** Representative bright field and fluorescence images of the mCTC *in vitro* cell culture. mCTC cells express eGFP (Scale bar 100 µm). **D** Representative histograms of cell cycle analysis of MDA-MB-231, MDA-MB-231-ULA, and mCTC cells (n=2). **E** Schematic representation of the zebrafish embryo metastasis assay and dot plot showing the fluorescence intensity of disseminated mCTC and MDA-MB-231-ULA cells in the tail of the zebrafish at 24 and 72 hours post-injection (n= 3). $^{**}p < 0.01$, $^{****}p < 0.0001$. **F** Representative images of single and clustered mCTC cells microinjected into the zebrafish embryo. Red arrows point towards the cells (scale bars 250 µm).

**Supplementary Figure 2. mCTC RNA-seq analysis and hit validation. A** Gene enrichment analysis of DEGs per biological process, cellular component and molecular function. **B** Relative mRNA expression of *SPARC*, *COL6A3*, *ITGB3*, and *TIMP3* observed in CAFs, mCTC, MDA-MB-231, and MDA-MB-231-ULA. Data are expressed relative to the average expression levels of β-2-microglobulin (β2M), which was used as the housekeeping gene (n=1). C mRNA expression level of *SPARC* in colorectal cancer (CCR), head and neck squamous cell carcinoma (HNSC), pancreatic adenocarcinoma (PDC), and melanoma (MEL). $^{*}p < 0.05$. **D** Kaplan-Meier plot of OS analysis according to *SPARC* expression levels in the BRCA TCGA dataset. Breast cancer patients were divided in two subgroups according to the high or low expression of *SPARC*.

**Supplementary Figure 3. Gene expression of genes dysregulated in CTC-clusters and prognostic value. A** Heatmap showing the expression levels in single CTCs and CTC-clusters of the 14 genes identified by the PPI analysis (GSE51827 and GSE86978 datasets). **B** mRNA expression level of *THBS1*, *VCL*, and *HSP90AB1* in the TCGA database (GEPIA analysis). **C** Kaplan-Meier plot of OS analysis according to the expression levels of *THBS1*, *VCL*, and *HSP90AB1* in the BRCA TCGA dataset. Patients were divided in two subgroups according to the high or low expression of the genes.

**Supplementary Figure 4. Batch effect correction of microarray data of the validation cohort and TCGA xCell analysis. A** Principal component analysis (PCA) of mRNA gene expression data before and after batch effect correction of the 11 datasets included in the validation cohort. **B** Box plots showing the different infiltrating abundances of immune and stromal cells in the TCGA dataset based on the classification of patient risk groups according to the CTCc-gene signature (xCell analysis).

**Supplementary Fig. S1**

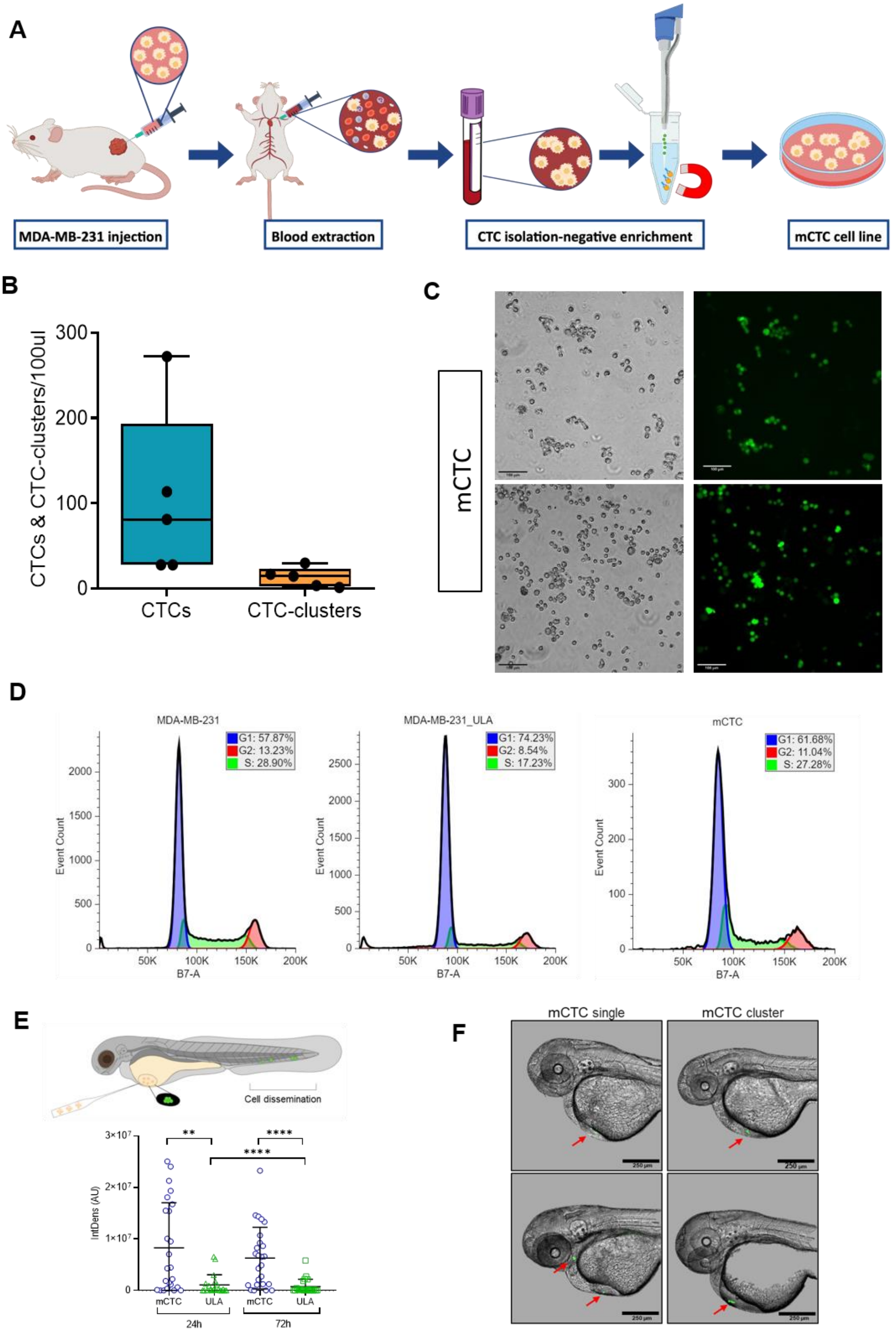

### Supplementary Fig. S2

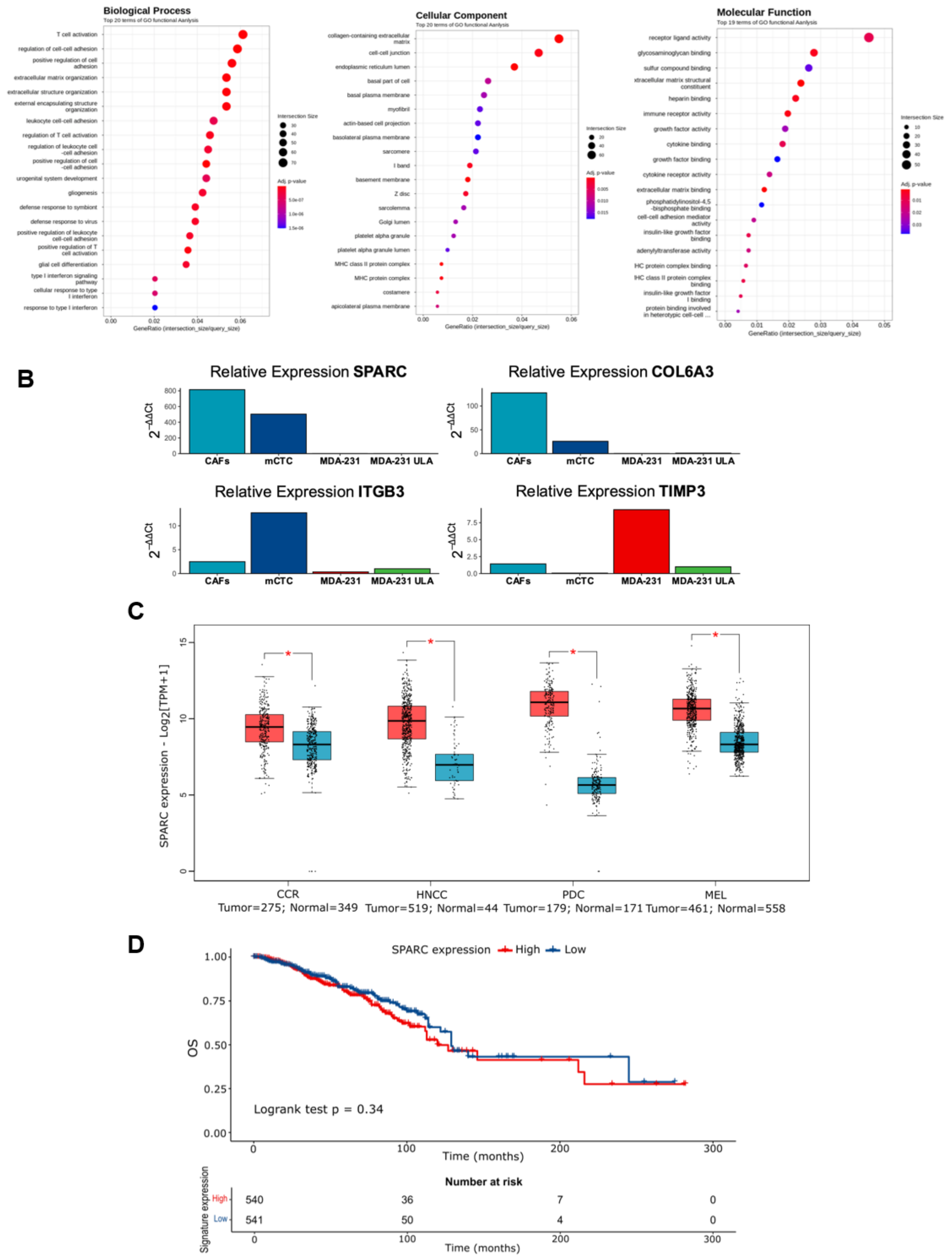

**Supplementary Fig. S3**

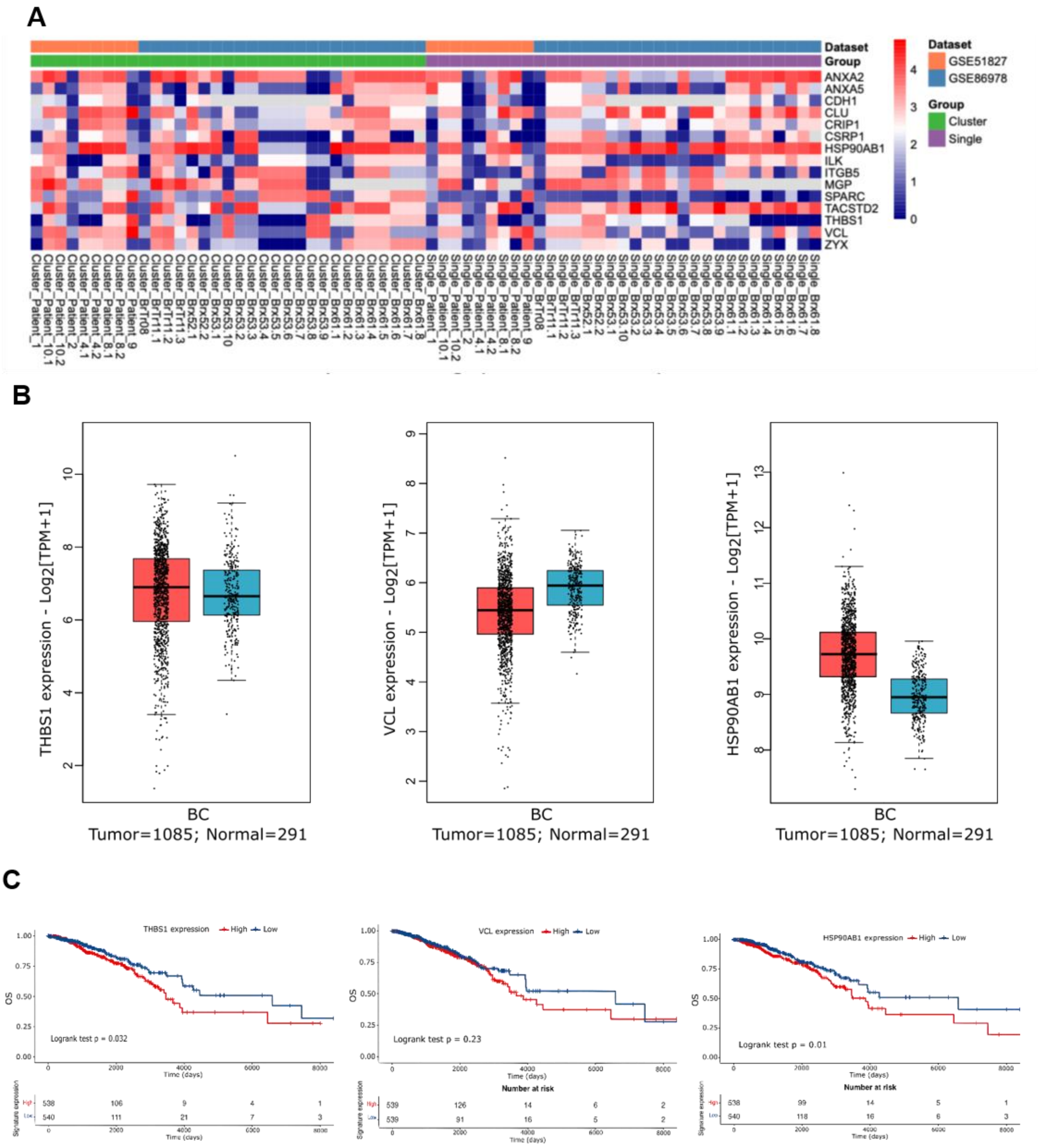

**Supplementary Fig. S4**

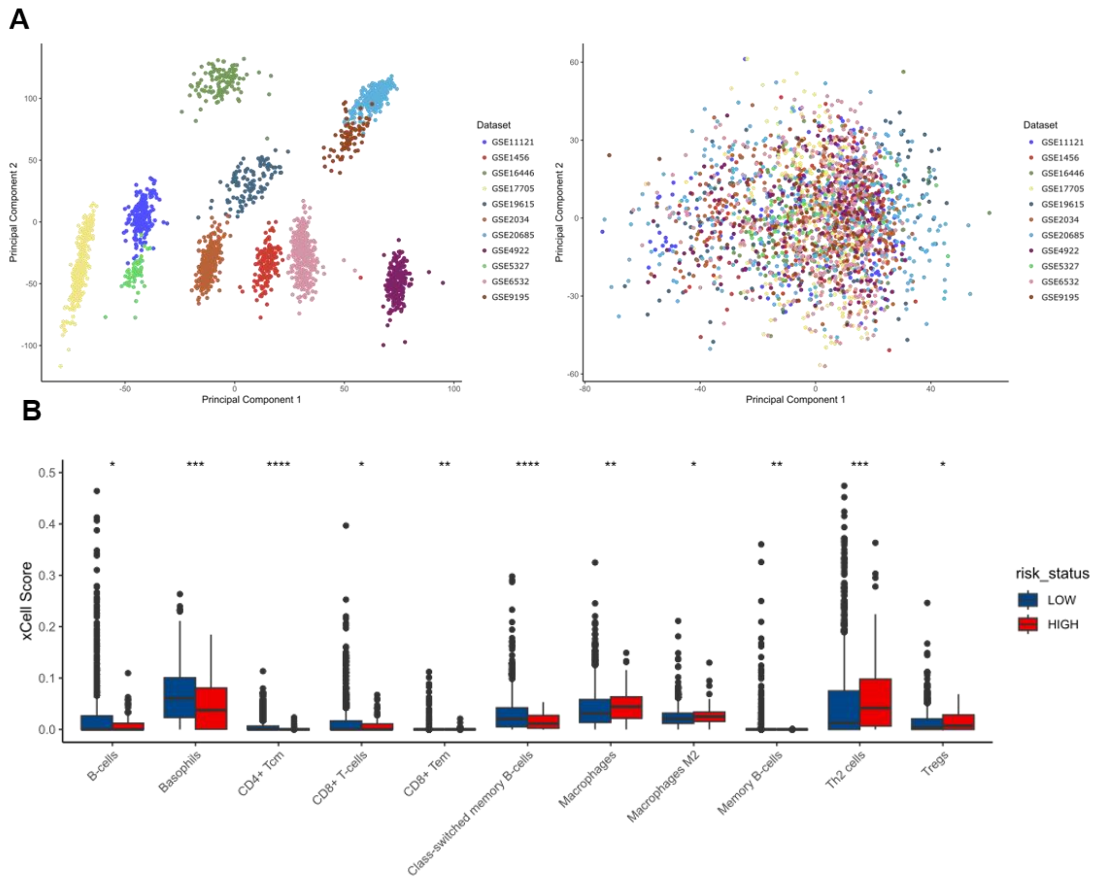
